# Analytical performance of rapid antigen tests for the detection of SARS-CoV-2 during widespread circulation of the Omicron variant

**DOI:** 10.1101/2022.05.17.22275034

**Authors:** Hiromichi Suzuki, Yusaku Akashi, Daisuke Kato, Yuto Takeuchi, Yoshihiko Kiyasu, Norihiko Terada, Yoko Kurihara, Miwa Kuwahara, Shino Muramatsu, Atsuo Ueda, Shigeyuki Notake, Koji Nakamura

## Abstract

**Introduction:** Antigen testing is essential in the clinical management of COVID-19. However, most evaluations of antigen tests have been performed before the emergence of the Omicron variant. Thus, an assessment of the diagnostic performance of antigen tests for the detection of SARS-CoV-2 during the circulation of Omicron variant is required.

**Methods:** This prospective observational study evaluated QuickNavi-COVID19 Ag, a rapid antigen detection test between December 2021 and February 2022 in Japan, using real-time reverse transcription (RT)-PCR as a reference. Two nasopharyngeal samples were simultaneously collected for antigen testing and for RT-PCR. Variant analysis of the SARS-CoV-2 genomic sequencing was also performed.

**Results:** In total, nasopharyngeal samples were collected from 1,073 participants (417 positive; 919 symptomatic; 154 asymptomatic) for analysis. Compared with those of RT-PCR, the sensitivity, specificity, positive predictive value, and negative predictive value were 94.2% (95% CI: 91.6%–96.3%), 99.5% (95% CI: 98.7%–99.9%), 99.2% (95% CI: 97.8%–99.8%), and 96.5% (95% CI: 94.8%–97.7%), respectively. The sensitivity among symptomatic individuals was 94.3% (95% CI: 91.5%–96.4%). Overall, 85.9% of sequences were classified as Omicron sublineage BA.1, 12.4% were Omicron sublineage BA.2, and 1.6% were Delta B.1.617.2. (Delta variant). Most of the samples (87.1%) had Ct values <25.

**Conclusions:** The QuickNavi-COVID19 Ag test showed high diagnostic performance for the detection of the SARS-CoV-2 Omicron sublineages BA.1 and BA.2 from nasopharyngeal samples.

## Introduction

The emergence of severe acute respiratory syndrome coronavirus 2 (SARS-CoV-2) B.1.1.529, i.e., the Omicron variant, dramatically increased the clinical cases of coronavirus disease 2019 (COVID-19). Owing to its high transmissibility, short incubation period [1-4] and reduced vaccine efficacy [5,6], the Omicron variant sublineages BA.1 and BA.2 became predominant worldwide by early 2022 [7]. Compared with previously dominant variants, COVID-19 caused by the Omicron variant is less likely to damage the lung [8] and more frequently causes sore throat and a hoarse voice [9].

Qualitative antigen tests that use immunochromatography are a useful point-of-care diagnostic testing method for infectious diseases because of their low cost, simple procedure, high availability of the test device, and short analytical time. For the laboratory diagnosis of COVID-19, antigen testing has been recommended for both symptomatic and asymptomatic individuals who are at high risk of infection, especially in situations where the prevalence of SARS-CoV-2 is ≥5% or where nucleic acid amplification test capacity is limited [10]. The diagnostic performance of antigen testing had been presumed to be preserved for the Omicron variant [7]. However, a significant impairment of sensitivity to the Omicron variant has been reported for nine antigen tests [11,12].

Herein, we prospectively evaluated the diagnostic performance of a qualitative antigen test (QuickNavi-COVID19 Ag, Denka Co., Ltd., Tokyo, Japan) using nasopharyngeal swab samples. We also conducted a genomic sequencing analysis to identify SARS-CoV-2 variants.

## Patients and methods

This study was conducted between December 28, 2021 and February 16, 2022. Sample collection and antigen testing were performed at a drive-through sample collection point at Tsukuba Medical Center Hospital (TMCH), and PCR was performed in the TMCH microbiology department. TMCH provides SARS-CoV-2 testing for the Tsukuba district in Japan. People with and without symptoms were referred from 65 clinics and a local public health center. All asymptomatic individuals had a history of contact with a confirmed or suspected COVID-19 cases.

Informed consent was verbally obtained from all participants and was documented in their electronic medical record to prevent infection transmission, written informed consent was not obtained. The ethics board of the University of Tsukuba Hospital approved the study (approval number: R03-042), including the method of obtaining informed consent.

### Study process

Two nasopharyngeal samples were separately collected by medical professionals one for RT-PCR and the other for antigen testing, as previously described [13-20]. A nasopharyngeal sample was obtained from each nasal cavity. All antigen tests were immediately performed on site after sample collection. A swab was inserted into a specimen buffer tube, and three drops of the prepared specimen were added on the test device. The sample processing time was 8 minutes, and the result was analyzed visually by the personnel who collected the sample.

For RT-PCR, a swab was diluted in 3 mL of Universal Transport Medium (Copan Italia S.p.A., Brescia, Italy) on site, and the sample was transferred to the TMCH microbiology department for in-house RT-PCR testing [13,20]. A 200 μL aliquot of each nasopharyngeal sample was extracted with a magLEAD 6gC (Precision System Science Co., Ltd., Chiba, Japan), and 100 μL of purified sample was eluted. The eluted samples were transferred to Denka Co., Ltd. For reference real-time RT-PCR testing to identify SARS-CoV-2, we used a method developed by the National Institute of Infectious Diseases (NIID), Japan. This method used an Applied Biosystems QuantStudio 3 (Thermo Fisher Scientific Inc., Waltham, MA, USA) with a QuantiTect probe RT-PCR kit (Qiagen Inc., Germantown, MD, USA) and a primer/probe N and N2 set [22]. Until the evaluation, all samples were preserved at −80 °C.

In case of discrepancy between the in-house PCR and the NIID method results for the presence or absence of SARS-CoV-2, additional examinations with the Xpert Xpress SARS-CoV-2 and GeneXpert system (Cepheid, Sunnyvale, CA, USA) [23] were performed, and those results were used as the final judgment.

### SARS-CoV-2 variant analysis

Of the 393 RT-PCR and QuickNavi-COVID19 Ag positive samples, 185 samples with high viral load (Ct ≤ 21) were subjected to genomic sequencing analysis. RNAs of 185 samples were extracted using QIAGEN Viral RNA mini kit and sent to Denka Co., Ltd. Denka Co., Ltd. then sent the RNA to iLAC Inc. (Ibaraki, Japan) requesting to perform Next-Generation Sequencing (NGS) and the SARS-CoV-2 variant analysis. NGS was performed using Illumina’s COVIDSeq test and IDT for Illumina-PCR indexes Sets 1-4 (Illumina, San Diego, CA, USA) to prepare sequencing libraries. Sequencing runs were performed on the prepared libraries using Illumina NovaSeq 6000 sequencer and NovaSeq 6000 SP Reagent Kit ver1.5. Sequencing results were then analyzed for SARS-CoV-2 variants using Illumina’s DRAGEN COVIDSeq Test Pipeline.

Specificity evaluation of QuickNavi-COVID19 Ag and QuickNavi-Flu+COVID19 Ag We also evaluated eight different lots of QuickNavi-COVID19 Ag and QuickNavi-Flu+COVID19 Ag tests, including products that were within 3 months of the expiration date (QuickNavi-COVID19 Ag lot numbers: 0700121, 1051081, 1071091, 1221101, 1231101 and 1241101, QuickNavi-Flu+COVID19 Ag, lot numbers: 0081081 and 0151091). The two test types were carried out with the same study processes. This supplemental evaluation was conducted during a non-surge period of COVID-19 (between November, 2021 and January, 2022) to estimate the false-positive rate for SARS-CoV-2 detection. For the evaluation, three test devices were simultaneously evaluated with one prepared specimen.

### Statistical analyses of the rapid antigen test

The sensitivity, specificity, positive predictive value (PPV), and negative predictive value (NPV) of the antigen tests were calculated with 95% confidence intervals (CIs).

The sensitivity stratified by the cycle threshold (Ct) value based on the N2 set of the NIID method was also evaluated. All statistical analyses were conducted using R 4.1.2 software (R Foundation, Vienna, Austria) with the “readxl,” “tidyverse,” and “epiR” packages.

## Results

Nasopharyngeal samples were collected from 1,073 participants during the study period; 919 were from symptomatic individuals and 154 were from asymptomatic individuals. For symptomatic participants, the median duration from symptom onset to sample collection was 2 days (interquartile range: 1–3 days).

Of the 1,073 samples, 411 were SARS-CoV-2 positive by RT-PCR with the NIID method. There were six discordances between the NIID and in-house RT-PCR results. Of the six discordant samples, all were SARS-CoV-2 positive when analyzed by the GeneXpert® system. Consequently, 417 samples (38.9%) were considered to be positive.

The QuickNavi-COVID19 Ag results are shown in Table 1. The sensitivity, specificity, PPV, and NPV were 94.2% (95% CI: 91.6%–96.3%), 99.5% (95% CI: 98.7%–99.9%), 99.2% (95% CI: 97.8%–99.8%), and 96.5% (95% CI: 94.8%–97.7%), respectively. For symptomatic individuals (Table 2a), the sensitivity was 94.3% (95% CI:91.5%-96.4%) and the specificity was 99.8% (95% CI: 99.0%–100%). For asymptomatic individuals (Table 2b), the sensitivity was 93.1% (95% CI: 77.2%–99.2%) and the specificity was 98.4% (95% CI: 94.3%–99.8%).

**Table 1.** Sensitivity and specificity of the QuickNavi-COVID19 Ag test among all samples

|  |  | real-time RT-PCR |  |
| --- | --- | --- | --- |
|  |  | Positive | Negative |
| Antigen test | Positive | 393 | 3 |
|  | Negative | 24 | 653 |
| Sensitivity (%) |  | 94.2 (91.6 - 96.3) |  |
| Specificity (%) |  | 99.5 (98.7 - 99.9) |  |

RT-PCR, reverse transcription-polymerase chain reaction
Data in parentheses indicate 95% confidence intervals.

**Table 2a.** Sensitivity and specificity of the QuickNavi-COVID19 Ag test among symptomatic individuals

|  |  | real-time RT-PCR |  |
| --- | --- | --- | --- |
|  |  | Positive | Negative |
| Antigen test | Positive | 366 | 1 |
|  | Negative | 22 | 530 |
| Sensitivity (%) |  | 94.3 (91.5 – 96.4) |  |
| Specificity (%) |  | 99.8 (99.0 – 100) |  |

RT-PCR, reverse transcription-polymerase chain reaction
Data in parentheses indicate 95% confidence intervals.

**Table 2b.** Sensitivity and specificity of the QuickNavi-COVID19 Ag test among asymptomatic individuals

|  |  | real-time RT-PCR |  |
| --- | --- | --- | --- |
|  |  | Positive | Negative |
| Antigen test | Positive | 27 | 2 |
|  | Negative | 2 | 123 |
| Sensitivity (%) |  | 93.1(77.2 - 99.2) |  |
| Specificity (%) |  | 98.4(94.3 – 99.8) |  |

RT-PCR, reverse transcription-polymerase chain reaction
Data in parentheses indicate 95% confidence intervals.

The antigen test sensitivities stratified by Ct value (N2) are shown in Table 3. For Ct values of <20, 20–24, 25–29, and ≥30, the sensitivities were 98.9% (95% CI: 96.1%–99.9%), 97.8% (95% CI: 94.4%–99.4%), 85.7% (95% CI: 67.3%–96.0%), and 47.4% (95% CI: 24.4%–71.1%), respectively.

**Table 3.** Sensitivity of QuickNavi-COVID19 Ag stratified by Ct values

| Ct values (N2) | Sensitivity (%) | Positive | Negative |
| --- | --- | --- | --- |
| <20 | 98.9 (96.1- 99.9) | 181 | 2 |
| 20-24 | 97.8 (94.4- 99.4) | 177 | 4 |
| 25-29 | 85.7 (67.3- 96.0) | 24 | 4 |
| ≥30 | 47.4 (24.4 – 71.1) | 9 | 10 |

Ct, cycle threshold
Data in parentheses indicate 95% confidence intervals.

Table 4 summarizes the QuickNavi-COVID19 Ag and QuickNavi-Flu+COVID19 Ag false-positive rates during the non-surge period of COVID-19. No positive results were obtained by RT-PCR for this evaluation. In total, 1,200 tests were performed using 400 nasopharyngeal samples, but no false-positive rapid antigen test results were observed.

**Table 4.** Specificity of QuickNavi-COVID19 Ag and QuickNavi-Flu+COVID19 Ag during a non-surge period of COVID-19

| Product | Lot number | Expiration dates | Number of tests | Number of false positives |
| --- | --- | --- | --- | --- |
| QuickNavi-COVID19 Ag | 0700121 | 2021.12 | 200 | 0 |
|  | 1051081 | 2022.08 | 100 | 0 |
|  | 1071091 | 2022.09 | 100 | 0 |
|  | 1221101 | 2022.10 | 200 | 0 |
|  | 1231101 | 2022.10 | 200 | 0 |
|  | 1241101 | 2022.10 | 200 | 0 |
| QuickNavi-Flu+COVID19 Ag | 0081081 | 2022.08 | 100 | 0 |
|  | 0151091 | 2022.09 | 100 | 0 |
| Total |  |  | 1200 | 0 |

The SARS-CoV-2 genome analysis results are shown in Figure 1. Of the 185 samples, 140 (75.7%) were determined to be BA.1.1.2, 21 (11.4%) were BA.2.3, 13 (7.0%) were BA.1.1, 3 (1.6%) were BA.1., 3 (1.6%) were AY.29, 2 (1.1%) were BA.1.1.1, 2 (1.1%) were BA.2 and 1(0.5%) was BA.1.15. Overall, 85.9% of the samples were classified as Omicron variant sublineage BA.1, 12.4% were Omicron variant sublineage BA.2, and 1.6% were Delta variant B.1.617.2.

**Figure 1.**
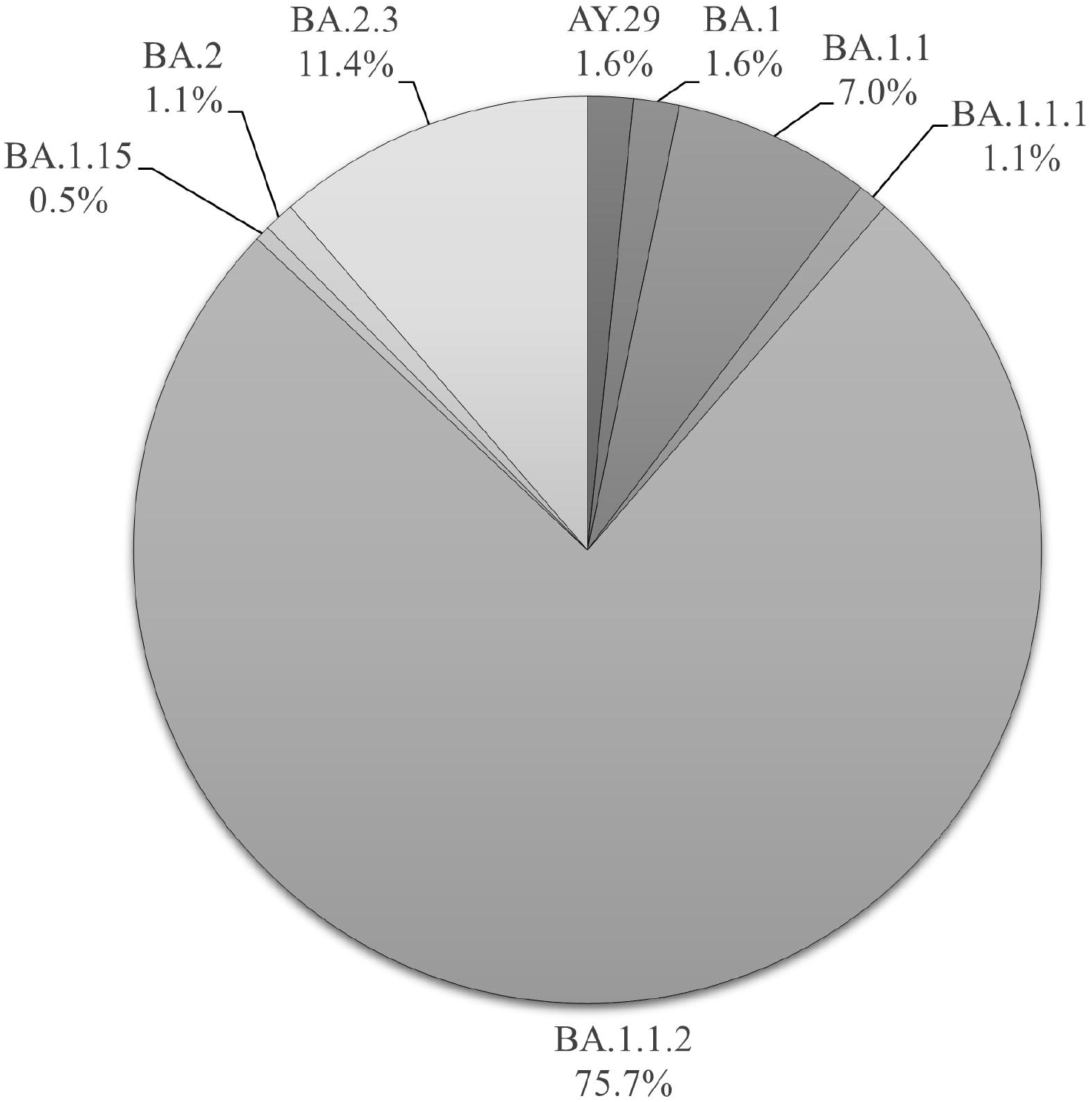
Proportions of SARS-CoV-2 variants by genomic sequencing analysis

## Discussion

In this study, more than 90% of individuals who were PCR positive for SARS-CoV-2 were correctly identified by rapid antigen testing, which included both symptomatic and asymptomatic individuals. However, the small number of asymptomatic participants might limit the generalizability of our results. Nevertheless, the current investigation showed that the QuickNavi-COVID19 Ag test has sufficient sensitivity for the detection of Omicron BA.1 and BA.2 from nasopharyngeal samples.

There has been insufficient investigation into the diagnostic performance of antigen testing for the Omicron variant. Osterman et al. demonstrated that nine SARS-CoV-2 antigen tests commercially available in Europe showed decreased sensitivities for the Omicron variant [12]. Bayart also showed that six antigen tests had significantly decreased sensitivities for samples with low viral loads of Omicron variant [11]. Meanwhile, the BinaxNOW (Abbott Diagnostics Scarborough Inc., ME, USA) rapid antigen test was shown to detect 95.2% (95% CI: 91%– 98%) of samples with RT-PCR Ct values <30 and 82.1% (95% CI: 77%–87%) of those with Ct values <35 during an Omicron surge period [24].

In a 2020–2021 clinical study, we evaluated the QuickNavi-COVID19 Ag test using 1,934 samples. The test sensitivity was 89.3% (95% CI: 82%–94%) for symptomatic individuals and 67.1% (95% CI: 55%–78%) for asymptomatic individuals, and no false positives were observed [15]. A re-evaluation of the QuickNavi-COVID19 Ag test was performed in 1,510 cases during the Delta variant dominantly circulating period in 2021, and the test sensitivity was 88.3% (95% CI: 83%–93%) for symptomatic individuals and 69.4% (95% CI: 60%–78%) for asymptomatic individuals. Three false-positive tests (0.2%) were identified (20). The current study found a slightly better clinical performance for the QuickNavi-COVID19 Ag test during the widespread Omicron circulating period. The improvement could be owing to the higher proportions of symptomatic individuals and people with high viral loads. The Ct values of people who are infected with the Omicron variant are considered to be nearly the same as for previous variants; thus, it is unclear why a high proportion of participants had a high viral load [25].

An increased false-positive rate has been reported for several lots of the QuickNavi-COVID19 Ag test due to inappropriate materials [26]; this problem was identified several months after production of the test kits. In this study, we performed 1,200 tests (eight lots) including the lots that were within 3 months of the expiration date, and no false positives were observed. In addition, there were only three false-positive samples identified during the Omicron variant period evaluation. Swabs for the antigen test and RT-PCR were obtained separately, and the sample collection procedure for the reference RT-PCR test may have been inappropriate. The flawed procedure may have caused false negatives, which was observed in a previous evaluation [20]. Clinicians should be aware that defective products are a possibility, especially if there are many positive results during a low-prevalence period.

This study has some limitations. First, the samples were collected at one site in Japan, and most samples were collected soon after symptom onset. The sample size for asymptomatic individuals might have been insufficient. Second, the assessment of lateral flow device results can vary among examiners [27]. Third, the reference RT-PCR examinations were performed with frozen samples, and the storage and transportation processes may have affected the test results. In addition, study samples were collected from the nasopharyngeal tract, and anterior nasal samples were not analyzed.

In conclusion, the current study showed that the QuickNavi-COVID19 Ag test had a high diagnostic performance for the detection of SARS-CoV-2 Omicron sublineages BA.1 and BA.2 in nasopharyngeal samples.

## Data Availability

All data produced in the present study are available upon reasonable request to the authors

## Acknowledgments

We thank Ms. Yoko Ueda, Ms. Mio Matsumoto, Mr. Masaomi Matsubayashi, Ms. Mika Yaguchi, Ms. Yumiko Tanaka, Mr. Naoki Tanimura, and the staff of the Department of Clinical Laboratory of Tsukuba Medical Center Hospital for their support in this study. We thank all participating medical institutions for providing their patients’ clinical information.

## Conflict of Interest

Denka Co., Ltd. provided funds for research expenses and the QuickNavi-Flu+COVID19 Ag and QuickNavi-COVID19 Ag tests without charge. Hiromichi Suzuki received a lecture fee from Otsuka Pharmaceutical Co., Ltd. Daisuke Kato, Miwa Kuwahara and Shino Muramatsu work for Denka Co., Ltd., the developer of the QuickNavi-Flu+COVID19 Ag and QuickNavi-COVID19 Ag tests.

## Notes

### Author Declarations

The ethics board of the University of Tsukuba Hospital approved the study (approval number: R03-042), including the method of obtaining informed consent.

## References

1. Backer JA, Eggink D, Andeweg SP, Veldhuijzen IK, van Maarseveen N, Vermaas K, et al. Shorter serial intervals in SARS-CoV-2 cases with Omicron BA.1 variant compared with Delta variant, the Netherlands, 2021. Euro Surveill. 2022;27.13-26

2. Cheng VC, Ip JD, Chu AW, Tam AR, Chan WM, Abdullah SMU, et al. Rapid spread of SARS-CoV-2 Omicron subvariant BA.2 in a single-source community outbreak. Clin Infect Dis 2022.(ahead of print)

3. Lee HR, Choe YJ, Jang EJ, Kim J, Lee JJ, Lee HY, et al. Time from Exposure to Diagnosis among Quarantined Close Contacts of SARS-CoV-2 Omicron Variant Index Case-Patients, South Korea. Emerg Infect Dis 2022;28:901–3.

4. Viana R, Moyo S, Amoako DG, Tegally H, Scheepers C, Althaus CL, et al. Rapid epidemic expansion of the SARS-CoV-2 Omicron variant in southern Africa. Nature 2022;603:679–86.

5. Andrews N, Stowe J, Kirsebom F, Toffa S, Rickeard T, Gallagher E, et al. Covid-19 Vaccine Effectiveness against the Omicron (B.1.1.529) Variant. N Engl J Med 2022;386:1532–46.

6. Tseng HF, Ackerson BK, Luo Y, Sy LS, Talarico CA, Tian Y, et al. Effectiveness of mRNA-1273 against SARS-CoV-2 Omicron and Delta variants. Nat Med. 2022. (ahead of print)

7. World Health Organization. Weekly epidemiological update on COVID-19 - 27 April 2022. https://www.who.int/publications/m/item/weekly-epidemiological-update-on-covid-19---27-april-2022 [Accessed 9 May 2022]

8. Hui KPY, Ho JCW, Cheung MC, Ng KC, Ching RHH, Lai KL, et al. SARS-CoV-2 Omicron variant replication in human bronchus and lung ex vivo. Nature 2022;603:715–20.

9. Menni C, Valdes AM, Polidori L, Antonelli M, Penamakuri S, Nogal A, et al. Symptom prevalence, duration, and risk of hospital admission in individuals infected with SARS-CoV-2 during periods of omicron and delta variant dominance: a prospective observational study from the ZOE COVID Study. Lancet 2022;399:1618–24.

10. World Health Organization. Antigen-detection in the diagnosis of SARS-CoV-2 infection using rapid immunoassays. https://www.who.int/publications/i/item/antigen-detection-in-the-diagnosis-of-sars-cov-2infection-using-rapid-immunoassays

11. Bayart JL, Degosserie J, Favresse J, Gillot C, Didembourg M, Djokoto HP, et al. Analytical Sensitivity of Six SARS-CoV-2 Rapid Antigen Tests for Omicron versus Delta Variant. Viruses 2022;14 :654.

12. Osterman A, Badell I, Basara E, Stern M, Kriesel F, Eletreby M, et al. Impaired detection of omicron by SARS-CoV-2 rapid antigen tests. Med Microbiol Immunol. 2022. (Ahead of print)

13. Kiyasu Y, Akashi Y, Sugiyama A, Takeuchi Y, Notake S, Naito A, et al. A Prospective Evaluation of the Analytical Performance of GENECUBE((R)) HQ SARS-CoV-2 and GENECUBE((R)) FLU A/B. Mol Diagn Ther 2021;25:495–504.

14. Kiyasu Y, Owaku M, Akashi Y, Takeuchi Y, Narahara K, Mori S, et al. Clinical evaluation of the rapid nucleic acid amplification point-of-care test (Smart Gene SARS-CoV-2) in the analysis of nasopharyngeal and anterior nasal samples. J Infect Chemother 2022;28:543–7.

15. Kiyasu Y, Takeuchi Y, Akashi Y, Kato D, Kuwahara M, Muramatsu S, et al. Prospective analytical performance evaluation of the QuickNavi-COVID19 Ag for asymptomatic individuals. J Infect Chemother 2021;27:1489–92.

16. Kurihara Y, Kiyasu Y, Akashi Y, Takeuchi Y, Narahara K, Mori S, et al. The evaluation of a novel digital immunochromatographic assay with silver amplification to detect SARS-CoV-2. J Infect Chemother 2021;27:1493–7.

17. Marty FM, Chen K, Verrill KA. How to Obtain a Nasopharyngeal Swab Specimen. N Engl J Med 2020;382:e76.

18. Suzuki H, Akashi Y, Ueda A, Kiyasu Y, Takeuchi Y, Maehara Y, et al. Diagnostic performance of a novel digital immunoassay (RapidTesta SARS-CoV-2): A prospective observational study with nasopharyngeal samples. J Infect Chemother 2022;28:78–81.

19. Takeuchi Y, Akashi Y, Kato D, Kuwahara M, Muramatsu S, Ueda A, et al. The evaluation of a newly developed antigen test (QuickNavi-COVID19 Ag) for SARS-CoV-2: A prospective observational study in Japan. J Infect Chemother 2021;27:890–4.

20. Takeuchi Y, Akashi Y, Kiyasu Y, Terada N, Kurihara Y, Kato D, et al. A prospective evaluation of diagnostic performance of a combo rapid antigen test QuickNavi-Flu+COVID19 Ag. J Infect Chemother 2022. (Ahead of print)

21. Naito A, Kiyasu Y, Akashi Y, Sugiyama A, Michibuchi M, Takeuchi Y, et al. The evaluation of the utility of the GENECUBE HQ SARS-CoV-2 for anterior nasal samples and saliva samples with a new rapid examination protocol. PLoS One. 2021;16:e0262159.

22. Shirato K, Nao N, Katano H, Takayama I, Saito S, Kato F, et al. Development of Genetic Diagnostic Methods for Detection for Novel Coronavirus 2019(nCoV-2019) in Japan. Jpn J Infect Dis. 2020;73:304–7.

23. Loeffelholz MJ, Alland D, Butler-Wu SM, Pandey U, Perno CF, Nava A, et al. Multicenter Evaluation of the Cepheid Xpert Xpress SARS-CoV-2 Test. J Clin Microbiol. 2020;58. e00926–20.

24. Schrom J, Marquez C, Pilarowski G, Wang CY, Mitchell A, Puccinelli R, et al. Comparison of SARS-CoV-2 Reverse Transcriptase Polymerase Chain Reaction and BinaxNOW Rapid Antigen Tests at a Community Site During an Omicron Surge : A Cross-Sectional Study. Ann Intern Med. 2022. (Ahead of print)

25. Laitman AM, Lieberman JA, Hoffman NG, Roychoudhury P, Mathias PC, Greninger AL. The SARS-CoV-2 Omicron Variant Does Not Have Higher Nasal Viral Loads Compared to the Delta Variant in Symptomatic and Asymptomatic Individuals. J Clin Microbiol. 2022;60:e0013922.

26. Denka Company Limited;. Second Notification. Notice of voluntary recall of certain lots of the COVID-19 rapid antigen test kit. https://www.denka.co.jp/eng/storage/news/pdf/377/20211115_denka_quicknavi_covid19ag_en.pdf

27. Peto T, Team UC-LFO. COVID-19: Rapid antigen detection for SARS-CoV-2 by lateral flow assay: A national systematic evaluation of sensitivity and specificity for mass-testing. EClinicalMedicine. 2021;36:100924.

